# Molecular characterization informs prognosis in patients with localized Ewing sarcoma: A report from the Children’s Oncology Group

**DOI:** 10.1101/2025.01.20.25320840

**Authors:** Riaz Gillani, David S. Shulman, Natalie J. DelRocco, Kelly Klega, Ruxu Han, Mark D. Krailo, Jonathan C. Slack, Mohammad Tanhaemami, Abigail Ward, Victoria Bainer, Cora Ricker, Josee Sparks, Kelly M. Bailey, Damon R. Reed, Steven G. DuBois, Patrick Leavey, Leo Mascarenhas, Patrick J. Grohar, Alanna J. Church, Brian D. Crompton, Katherine A. Janeway

**Author notes:** These authors contributed equally to this work (Riaz Gillani, David S. Shulman). These authors contributed equally to this work (Brian D. Crompton, Katherine A. Janeway).

## Abstract

**PURPOSE:** Identification of discrete sub-groups associated with treatment response and resistance in localized Ewing sarcoma (EWS) remains a challenge. The primary objective of the Children’s Oncology Group biology study AEWS18B1-Q was to perform molecular characterization of a large cohort of patients with localized Ewing sarcoma treated on prospective trials with modern standard of care therapy.

**METHODS:** We analyzed clinical and molecular features from patients with localized EWS enrolled on AEWS0031, AEWS1031, or INT-0154 frontline trials. All patients had available FFPE tissue, frozen tissue, or whole-genome amplified material. Sequencing was performed for identification of canonical fusions, recurrent copy number alterations (CNAs), and alterations in *TP53* and *STAG2*. Where available, tissue was analyzed for loss of STAG2 protein expression. Molecular features were evaluated for their association with cumulative incidence of relapse in univariate and multivariable analyses.

**RESULTS:** Three hundred and fifty-one cases had sufficient tissue, which in most cases was extracted from two FFPE slides. EWS canonical fusions were identified in 282 cases (80.3%). Pathogenic mutations in *TP53* and *STAG2* were identified in 5.1% and 7.6% of cases, respectively and 63.1% of cases were found to have recurrent CNAs. In univariate analysis, there was an increased cumulative incidence of relapse in patients with *TP53* mutation (5-year cumulative incidence of relapse 43%, CI [17%, 67%] vs. 22%, CI [17%, 27%]; Gray’s test *P* = 0.039), *STAG2* mutation (53%, CI [29%, 73%] vs. 21%, CI [16%, 26%]; *P* < 0.001), and recurrent CNAs (30%, CI [22%, 37%] vs. 16%, CI [9%, 24%]; *P* = 0.005). In a multivariable analysis, *STAG2* mutation was the only molecular biomarker that remained prognostic.

**CONCLUSION:** This is a prospective validation of the molecular prognostic features of localized EWS receiving standard of care therapy on therapeutic clinical trials. Building on prior work, patients with *STAG2* mutations were at high risk of relapse.

**CONTEXT:** *Key Objective:* To determine if molecular features can identify clinically relevant disease sub-groups applicable to frontline clinical trial design among patients with localized Ewing sarcoma.

*Knowledge Generated:* Among 351 patients with localized Ewing sarcoma treated on prospective trials, canonical fusions were identified in 80% of cases. In a multivariable analysis of patients with genomically defined Ewing sarcoma, the presence of *STAG2* mutations identified a high-risk population. Relevance (added by Associate Editor)

## INTRODUCTION

Ewing sarcoma (EWS) is an aggressive bone and soft tissue sarcoma defined by the presence of FET-ETS family fusions.^1^ The estimated 5-year event-free survival (EFS) is 78% for the approximately 70% of patients who present with localized disease enrolled on the most recent Children’s Oncology Group (COG) phase 3 trial with a treatment backbone of interval compressed (every 2 week) chemotherapy.^2^ Survivors are left with a large burden of late effects including cardiac dysfunction, second malignant neoplasms, infertility and physical disability.^3–5^ Given the relatively favorable survival outcomes with profound late morbidity in this population, there is an urgent need to define molecularly-characterized risk groups to inform approaches to risk-stratified therapy as has been done in the context of other solid tumors and hematologic malignancies,^6,7^ with the goal of improving cure rates for high-risk patients and reducing morbidity among patients with less aggressive disease.

The presence of metastatic disease is the strongest prognostic factor in EWS and clinical features alone have proven inadequate to further risk stratify patients with localized EWS.^8,9^ A growing body of evidence dating back to initial genomic landscape studies has contributed to our understanding of key molecular features of EWS.^10–12^ Prior studies have suggested that inactivation of *STAG2* and/or *TP53* may be associated with poor prognosis.^13,14^ Similarly, multiple recurrent copy number alterations (CNAs) have preliminary evidence for association with poor outcomes.^12,15,16^ These molecular biomarkers hold the potential to define prognostic sub-groups in EWS but require clinical validation in independent patient cohorts with prospective outcome data collection.^17^

Thus, given the emerging evidence for multiple prognostic molecular biomarkers in EWS, we pursued multimodal molecular analysis of a large cohort of patients with localized EWS treated with contemporary therapy on COG clinical trials. We sought to (a) search for disease-defining translocations, (b) describe the prevalence of relevant molecular biomarkers in a large cohort of patients with molecularly-defined localized EWS, and (c) identify molecularly-informed disease subgroups by testing for associations between molecular features (e.g., *STAG2* and *TP53* alterations) and outcomes.

## PATIENTS AND METHODS

### Study Population

Patients were required to have a pathologic diagnosis of EWS and be enrolled on and eligible for frontline clinical trials AEWS0031, AEWS1031, or INT-0154. Molecular confirmation of diagnosis was not required for trial eligibility. All patients were required to have newly diagnosed, localized EWS and available Formalin-Fixed Paraffin-Embedded (FFPE) tissue, frozen tissue, or previously sequenced whole-genome amplified (WGA) DNA from frozen tissue. Tissue was obtained from time of diagnosis in 95% of cases. The minimum quantity requested for FFPE material was two unstained slides. Among prospectively sourced patients enrolled to AEWS0031, preference was given to samples from patients who received interval compressed chemotherapy over non-interval compressed therapy. All patients signed informed consent at the time of enrollment to either AEWS0031, AEWS1031, or INT-0154. Separate approvals for this study were obtained from the Dana-Farber Cancer Institute Institutional Review Board, COG, and NCI.

### Sample Preparation and Sequencing

Fresh frozen tissue or FFPE was requested from the COG Biopathology Center (BPC), and DNA and RNA were extracted and quantified using standard methods. WGA DNA, generated from frozen tumor tissue for a previously published study, was available from a subset of patients treated on AEWS0031.^14^ Ultra-low passage whole genome sequencing (ULP-WGS) and the TranSS-Seq assay were run on all samples as previously described.^18,19^ Sequencing and computational methods are described in **Appendix A**.

### STAG2 Immunohistochemistry

STAG2 immunohistochemistry was performed using a previously described mouse anti-human monoclonal antibody and scored by two pediatric pathologists. H-scores were generated and cases with an H-score of 0 were categorized as having complete loss of STAG2 expression. Details of immunohistochemistry and scoring are described in **Appendix A**.

### Statistical Methods

For descriptive analyses, categorical variables were presented as counts (percents) and continuous variables were summarized by medians. Associations between two categorical variables used the X^2^ test or Fisher’s exact test if any cell counts were less than five. The distributions of continuous, skewed molecular analysis quality control variables were compared using the Wilcoxon Rank Sum test.

Post-enrollment cumulative incidence of relapse was the primary outcome measure for this study. Second malignant neoplasms and deaths as first events were treated as competing risks. For univariate analyses, cumulative incidence curves were plotted and compared where appropriate using Gray’s test for equality of cumulative incidence functions.^20^ Supplemental analyses were also conducted for event-free survival and overall survival using the Kaplan-Meier estimator and logrank test for equality of survival curves.

To estimate the association between molecular features and risk of relapse while controlling for clinical features, a multivariable Fine and Gray model for cumulative incidence of relapse was used.^21^ Clinical investigators defined predictors to be included in the model prior to analysis of outcome data. These were determined by clinical importance and presence of missing data. If a potential prognostic factor had more than 25% missingness, it was removed from consideration for inclusion in the final multivariable model. See **Appendix Table A2** for further details. This *a priori* variable selection approach was chosen due to the well-known limitations of stepwise variable selection and univariate screening.^22,23^

For the population of patients who received modern interval compressed chemotherapy and had molecularly defined EWS (i.e., the population that would be most similar to future trial populations), two different risk groupings were explored: (a) molecular and (b) multivariable model-derived. Three molecular subgroups defined (a): 1) patients with *STAG2* mutation, 2) patients with *TP53* mutation and/or recurrent CNAs but no *STAG2* mutation, and 3) patients with no identified molecular lesion. To visualize clinically meaningful risk groups from the multivariable model, which incorporates clinical risk factors as well as molecular, risk groups in (b) were based on target five-year EFS rates determined by clinical investigators as 90% for a low- and 50% for a high-risk group. Participants were ordered by their model-predicted risk of relapse and were sequentially added to low-risk and high-risk groups starting with ten patients (for estimation accuracy) until the target rate was exceeded. All remaining patients were considered to be intermediate-risk.

As a sensitivity analysis, all analyses were repeated using multiple imputation to account for missing data via the MICE algorithm.^24^ A two-sided *p*-value of ≤0.05 was considered significant in all analyses. No adjustment was made to account for the number of tests performed. All statistical analyses were performed using R Version 4.3.3 (R Core Team [2023]. R: A Language and Environment for Statistical Computing. R Foundation for Statistical Computing, Vienna, Austria. <https://www.R-project.org/>).

## RESULTS

### Patients and Samples

A total of 1,674 patients were enrolled on the parent trials AEWS0031, AEWS1031, and INT-0154. The initial analytic cohort included 354 unique patients with available FFPE (n=283), frozen tissue (n=6), or previously extracted WGA DNA (n=74; **Figure 1A**). Two cases were subsequently excluded due to trial ineligibility and the single case enrolled on INT-0154 was removed. 282 of the remaining 351 cases (80.3%) were found to have a canonical EWS fusion of *EWSR1-FLI1*, *EWSR1-ERG*, *EWSR1-ETV*, or *EWSR1-FEV* (**Figure 1B**). No *FUS* fusions were detected. Of note, fusions associated with other round cell sarcomas were identified in four cases (1.1%). The mean DNA content extracted from cases was 519 ng (range 16-5,565 ng).

**FIG 1.**
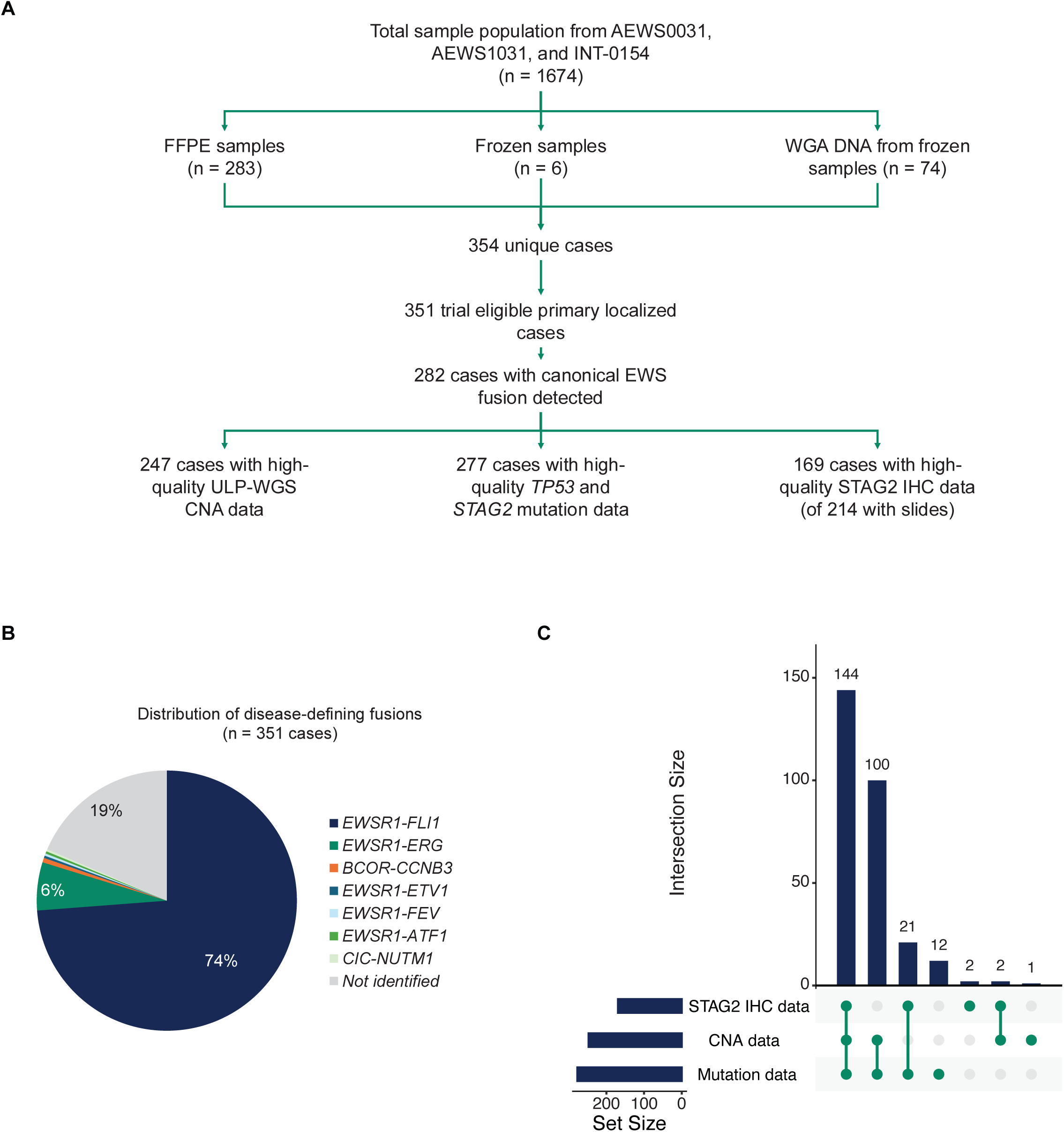
Cohort and study overview. (A) Consort diagram outlining source studies and case counts. Among 351 eligible cases with viable tissue, 282 cases were found to have canonical EWS fusions and included for further study. (B) The most common canonical EWS fusion identified was *EWSR1-FLI1* followed by *EWSR1-ERG*. 19% of cases had no fusion detected. (C) Large subsets of the analytic cohort were characterized by multiple molecular assays.

Cases with any detected fusion had higher overall DNA content (median 389.4ng for fusion positive cases vs. 141.5ng for fusion negative cases; Wilcoxon *P* < 0.001) and mean target coverage (median 717.5x for fusion positive cases vs. 11.2x for fusion negative cases; Wilcoxon *P* < 0.001) suggesting limitations in comprehensive fusion detection from archival tissue with minimal DNA content where only two FFPE slides were available for most cases. From the final analytic cohort of 282 patients with identified FET-ETS family fusions, 247 cases (87.6%) were evaluable for CNAs and 277 cases (98.2%) were evaluable for mutations. 214 cases had slides available for STAG2 IHC, and 169 cases (59.9%) had high-quality evaluable staining. Ninety-five percent of cases were represented with at least two molecular data modalities (**Figure 1C**).

In the final analytic cohort, 217 patients (77.0%) received treatment with interval compressed chemotherapy, 251 (89.0%) patients were less than 18 years of age at the time of trial enrollment and there was a slight male sex predominance, with 152 male patients (53.9%, **Table 1**). We observed no significant differences in clinical characteristics between the 282 patients with a canonical fusion in the analytic cohort and the 913 eligible patients who were not included, though a higher proportion of the analytic cohort was enrolled on AEWS1031.

**TABLE 1.** Study cohort clinical characteristics in comparison to unselected patients.

| Variable | Not included<br>(n = 913) | Included<br>(n = 282) | P value |
| --- | --- | --- | --- |
| Sex |  |  | 0.588 |
| Female | 402 (44.0%) | 130 (46.1%) |  |
| Male | 511 (56.0%) | 152 (53.9%) |  |
| Age |  |  | 0.059 |
| <18 years | 769 (84.2%) | 251 (89.0%) |  |
| ≥18 years | 144 (15.8%) | 31 (11.0%) |  |
| Race |  |  | 0.783 |
| American Indian or Alaska | 6 (0.7%) | 2 (0.7%) |  |
| Asian | 21 (2.3%) | 5 (1.8%) |  |
| Black or African American | 19 (2.1%) | 9 (3.2%) |  |
| Native Hawaiian or Other | 10 (1.1%) | 2 (0.7%) |  |
| White | 794 (87.0%) | 244 (86.5%) |  |
| NA | 63 (6.9%) | 20 (7.1%) |  |
| Ethnicity |  |  | >0.999 |
| Hispanic or Latino | 104 (11.4%) | 33 (11.7%) |  |
| Not Hispanic or Latino | 793 (86.9%) | 246 (87.2%) |  |
| NA | 16 (1.8%) | 3 (1.1%) |  |
| Tumor site |  |  | 0.639 |
| Extraosseous | 151 (16.5%) | 52 (18.4%) |  |
| Non-pelvic | 605 (66.3%) | 179 (63.5%) |  |
| Pelvic | 154 (16.9%) | 51 (18.1%) |  |
| NA | 3 (0.3%) | 0 (0.0%) |  |
| Tumor volume |  |  | 0.198 |
| <200 mL | 393 (43.0%) | 147 (52.1%) |  |
| ≥200 mL | 201 (22.0%) | 59 (20.9%) |  |
| NA | 319 (34.9%) | 76 (27.0%) |  |
| Interval compressed chemotherapy | 694 (76.0%) | 217 (77.0%) | 0.808 |
| Study |  |  | <0.001 |
| AEWS0031 | 462 (50.6%) | 105 (37.2%) |  |
| AEWS1031 | 451 (49.4%) | 177 (62.8%) |  |

### Univariate Analyses of Molecular Biomarkers

The 5-year cumulative incidence of relapse for the entire analytic cohort was 23.2% (95% CI [18.4%,28.4%]), with 5-year EFS of 73.5% (CI [68.4%,79.0%]) and 5-year OS of 81.4% (CI [76.9%,86.2%]) (**Appendix Figure A1**).

Among 282 patients with a detectable fusion, *EWSR1-FLI1* was the most common fusion, seen in 259 cases (91.8%), followed by *EWSR1-ERG*, seen in 21 cases (7.4%; **Figure 2A**). In a subset of 229 cases with *EWSR1-FLI1* fusions detected from DNA with unambiguous intronic breakpoints, Type I (57.2%) and Type II transcripts (24.0%) were most common (**Figure 2B**).^17,25^ There was no difference in cumulative incidence of relapse in patients with *EWSR1-FLI1* fusions vs. those with *EWSR1-ERG* fusions (23.6%, CI [18.5%,29.0%] vs. 20.6%, CI [6.1%,40.9%]; Gray‘s test *P* = 0.5; **Appendix Figure A2A**). Similarly, Type I *EWSR1-FLI1* fusions were not prognostic relative to all other *EWSR1-FLI1* fusion subtypes (27.6%, CI [20.0%,35.6%] vs. 21.3%, CI [13.6%,30.1%]; *P* = 0.2; **Appendix Figure A2B**).

**FIG 2.**
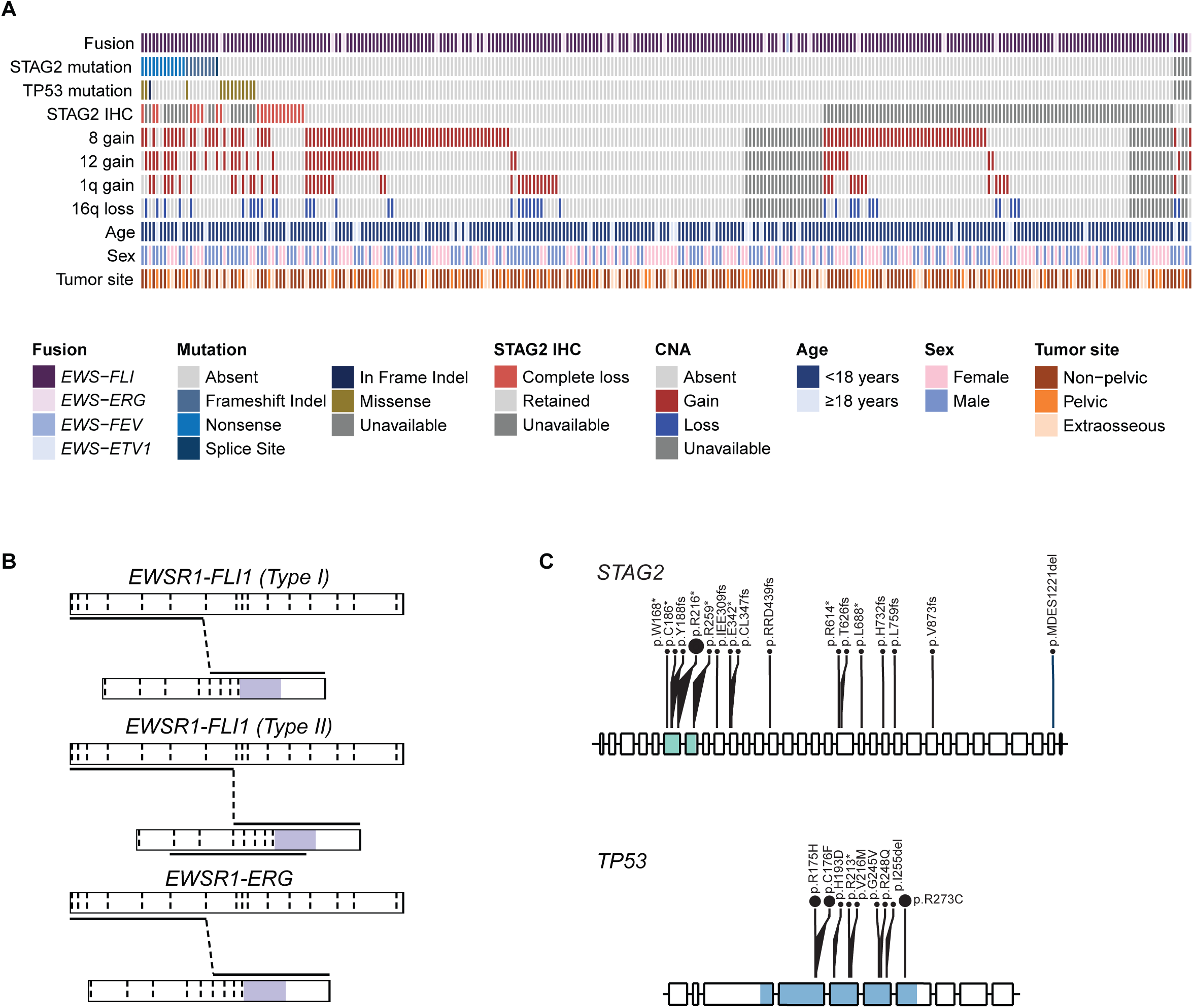
Molecular characterization of cohort. (A) Co-mutation plot summarizing molecular data across cohort of 282 primary localized EWS cases. (B) Illustrative examples of putative fusion transcripts of *EWSR1-FLI1* and *EWSR1-ERG* translocations observed in analytic cohort (purple = ETS domain). (C) Lollipop plots summarizing *STAG2* and *TP53* mutations in the analytic cohort (green = STAG domain, blue = P53 DNA-binding domain).

Pathogenic *TP53* and *STAG2* mutations were identified in 5.1% (14*/*277*) and* 7.6% (21/277) of patients, respectively. All the *STAG2* mutations were frameshift or nonsense mutations and the nonsense mutation p.R216* was recurrent. All *TP53* mutations were in the DNA binding domain and previously reported to be pathogenic. Several were recurrent, with *TP53* p.R273C being the most frequent (**Figure 2C**). Of the 247 cases evaluable for CNAs, there were 20.6% with chromosome 1q gain, 50.6% with chromosome 8 gain, 21.1% with chromosome 12 gain, and 17.0% with chromosome 16q loss. 169 cases were evaluable for STAG2 IHC, and complete STAG2 loss by IHC (normalized H score of 0) was seen in 22 (13%) cases (**Figure 2A**).

We assessed previously reported full chromosomal and arm-level CNAs as prognostic biomarkers (**Figure 3A**). While chromosome 1q gain trended toward a higher cumulative incidence of relapse (35.5%, CI [22.5%,48.6%] vs. 21.5%, [15.9%,27.7%]; *P* = 0.063), no single CNA reached statistical significance in univariate analyses (**Appendix Figure A3**). Patients with any recurrent CNA considered in aggregate (defined as any chromosome 1q gain, 8 gain, 12 gain, or 16q loss) had an increased cumulative incidence of relapse (29.6%, CI [22.4%, 37.1%] vs. 15.9%, CI [9.1%,24.3%]; *P* = 0.005; **Figure 3B**).

**FIG 3.**
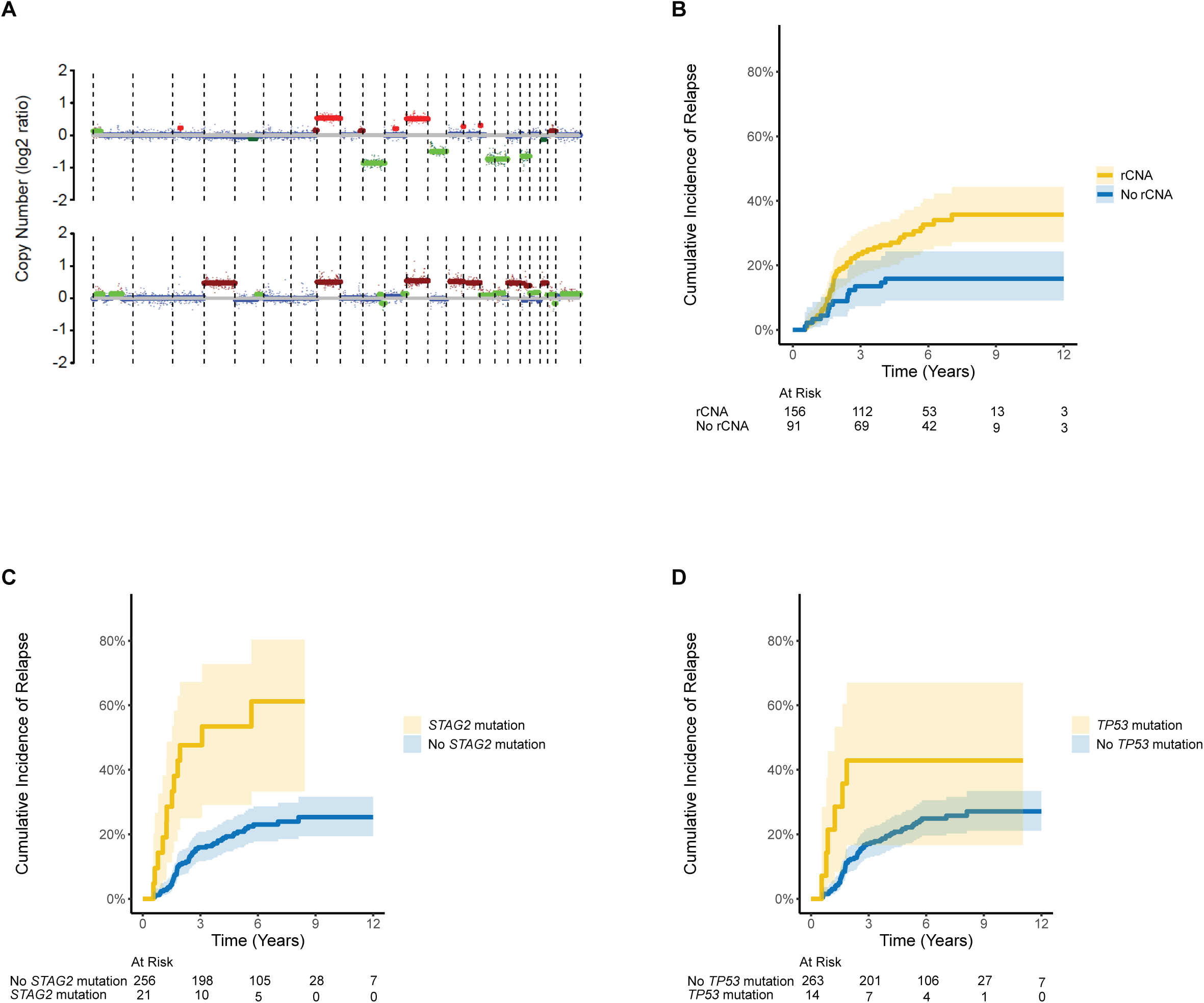
Cumulative incidence of relapse for recurrent CNAs, *STAG2* mutation, and *TP53* mutation. Chromosome 1q gain, chromosome 8 gain, chromosome 12 gain, and chromosome 16q loss were evaluated in this study using ichorCNA tracings derived from ULP-WGS, as represented in (A). Cumulative incidence of relapse for (B) recurrent CNAs (defined as any of the four CNAs evaluated), (C) *STAG2* mutation, and (D) *TP53* mutation.

The 5-year cumulative incidence of relapse in patients with a *STAG2* mutation was 53.4% (CI [29.1%,72.7%]), which was significantly higher than the cumulative incidence of relapse in patients without a *STAG2* mutation (20.8%, CI [15.9%,26.1%]; *P* < 0.001; **Figure 3C**). *TP53* mutations were also associated with a higher cumulative incidence of relapse (42.9%, CI [16.6%, 67.0%] vs. 22.2%, CI [17.2%,27.5%]; *P* = 0.039; **Figure 3D**).

EFS and OS analyses were also carried out for the preceding variables, demonstrating prognostic significance of *STAG2* mutation and *TP53* mutation using these outcome measures (**Appendix Figure A4**). Recurrent CNAs were not associated with inferior EFS or OS, potentially due to the presence of secondary malignancies among patients without recurrent CNAs in this cohort.

### Multivariable Model and Focused Analysis of Patients Receiving Interval Compressed Chemotherapy

We used a multivariable model to ascertain which features were most predictive of cumulative incidence of relapse when controlling for other factors. We observed that *STAG2* mutation (HR 3.52, CI [1.75, 7.10], *P* < 0.001) was the only molecular feature that remained prognostic in the multivariable analysis in addition to the clinical variables of age (HR 1.11, CI [1.06, 1.17], *P* < 0.001) and interval compressed chemotherapy (HR 0.37, CI [0.22, 0.63], *P* < 0.001) (**Table 2**). The same trends were observed when accounting for missing data using multiple imputation (**Appendix Table A3**).

**TABLE 2.** Multivariable analysis of association of molecular and clinical characteristics with cumulative incidence of relapse across 244 patients with complete data.

| Molecular/ clinical characteristic | HR | 95% CI | P value |
| --- | --- | --- | --- |
| rCNA | 1.79 | 0.96, 3.33 | 0.066 |
| TP53 mutation | 1.35 | 0.41, 4.48 | 0.6 |
| STAG2 mutation | 3.52 | 1.75, 7.10 | <0.001 |
| Interval compressed chemotherapy | 0.37 | 0.22, 0.63 | <0.001 |
| Age | 1.11 | 1.06, 1.17 | <0.001 |
| Male sex (relative to female sex) | 0.62 | 0.36, 1.04 | 0.072 |
| Site (relative to non-pelvic) |  |  |  |
| Extraosseous | 0.43 | 0.19, 1.02 | 0.054 |
| Pelvic | 0.95 | 0.53, 1.71 | 0.9 |

To see if we could use the high risk molecular and clinical features to identify prognostic risk groups, we used two approaches to risk stratify the patients included in the multivariable analysis who received interval compressed chemotherapy (n = 188). We defined three molecular subgroups: 1) patients with *STAG2* mutation, 2) patients with *TP53* mutation and/or recurrent CNAs but no *STAG2* mutation, and 3) patients with no identified molecular lesion beyond the fusion and determined EFS based on these groups. Using this approach, patients with *STAG2* mutation had increased risk of EFS-event when compared to patients with no identified molecular lesion (5-year EFS 51.4%, CI [30.8%,85.8%] vs. 80.3%, CI [71.2%,90.5%]; logrank *P* = 0.0014), but those with *TP53* mutation and/or recurrent CNAs without a *STAG2* mutation had a comparable risk of EFS-event (79.6%, CI [72.0%,88.1%]) to those with no molecular lesion (**Figure 4A**). We also used an outcome-based approach in the interval compressed chemotherapy cohort where low, intermediate, and high risk groups were predefined according to EFS (**Figure 4B**). In addition to a lower prevalence of *STAG2* mutations as established by the multivariable analysis, low-risk patients were characterized by younger age (mean age: 4.6 years in low-risk patients vs. 16.1 years in high-risk patients), smaller tumor volumes (proportion of evaluable tumors with volume ≥ 200 mL: 2/15 [13.3%] in low-risk patients vs. 11/26 [42.3%] in high-risk patients), and a higher proportion of extraosseous tumors (11/18 [61.1%] vs. 2/31 [6.5%]). Taken together, *STAG2* mutation showed prognostic value after controlling for receipt of interval compression and other clinical variables, and focused application of both risk group definitions to patients who received interval compressed chemotherapy confirmed that *STAG2* mutation was the driving molecular feature for identifying patients with high risk of relapse, while also suggesting that clinical variables may be important in identifying the patients with the lowest risk of relapse.

**FIG 4.**
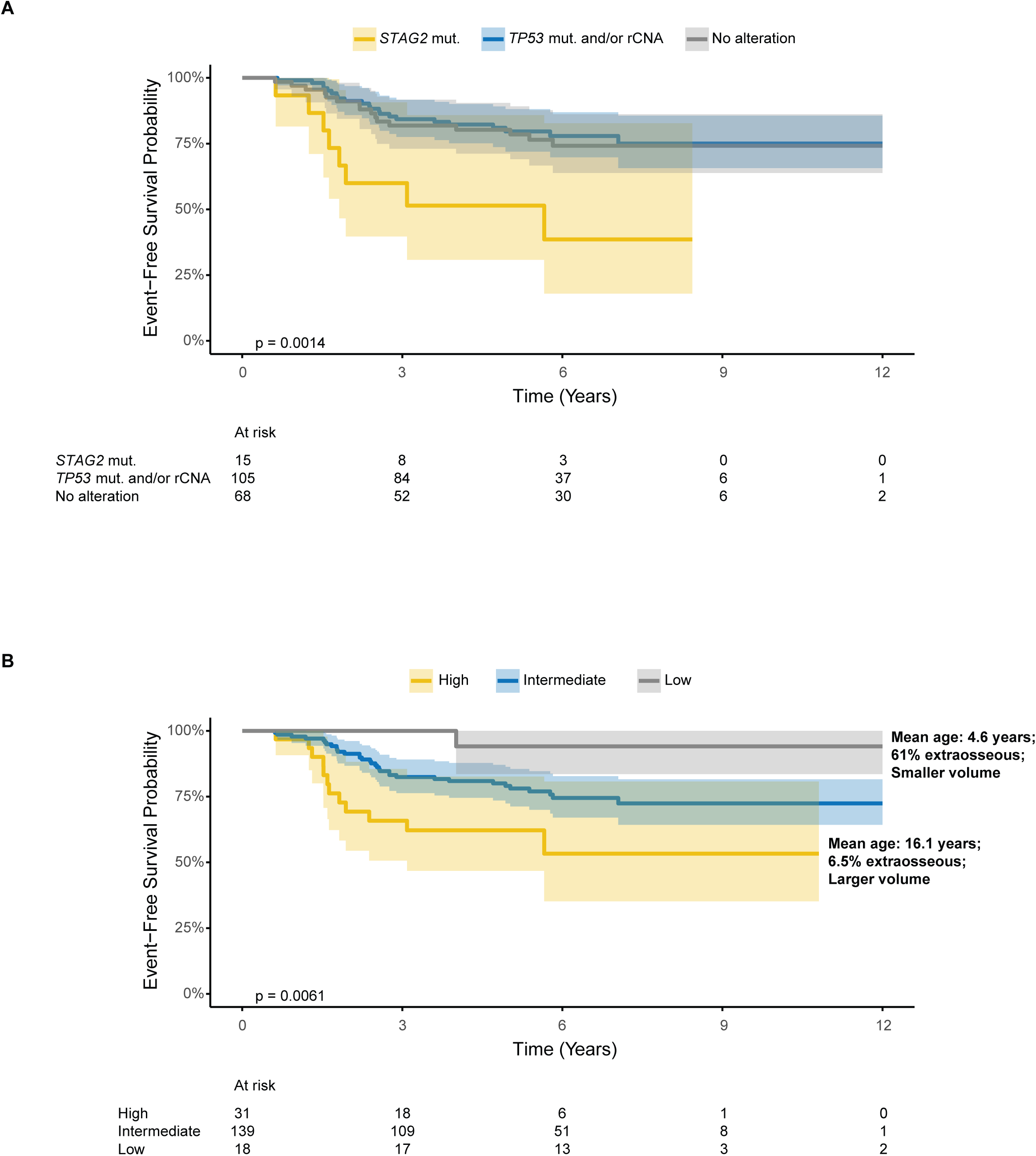
EFS when restricting only to patients who received interval compressed chemotherapy. (A) EFS for three molecularly characterized groups in this cohort: 1) patients with *STAG2* mutation, 2) patients with *TP53* mutation and/or recurrent CNAs but no *STAG2* mutation, and 3) patients with no identified molecular lesion. (B) When stratifying based on targeted EFS, low-risk patients were characterized by younger age, smaller tumor volumes, and a higher proportion of extraosseous tumors.

### Composite biomarker of STAG2 loss by mutation or IHC is associated with a poor outcome

Given the prior literature demonstrating that STAG2 loss of expression may occur with or without *STAG2* mutation,^11,13^ we evaluated STAG2 loss by IHC (**Appendix Figure A5A**) as well as the composite biomarker of STAG2 loss by mutation or IHC in univariate analyses. Where there were overlapping molecular data, we observed a high concordance between *STAG2* mutation and STAG2 loss by IHC, with 8 out of 10 *STAG2* mutations among cases ascertained to have complete STAG2 loss by IHC (odds ratio for STAG2 mutation 38.4, Fisher’s exact *P* < 0.001; **Appendix Figure A5B**). Conversely, 14 cases with STAG2 loss by IHC had no *STAG2* mutations, supporting alternative mechanisms of *STAG2* inactivation in addition to mutation.

STAG2 loss by IHC trended toward a higher cumulative incidence of relapse (32.6%, [14.1%,52.8%] vs. 17.4%, [11.5%,24.2%]; *P* = 0.075; **Appendix Figure A5C**), and the composite biomarker of STAG2 loss by mutation or IHC was associated with a higher cumulative incidence of relapse (38.1%, [21.9%,54.2%] vs. 16.6%, [11.0%,23.5%]; *P* = 0.001; **Appendix Figure A5D**).

## DISCUSSION

We show in a large cohort of patients with newly diagnosed, molecularly-defined, localized EWS treated with contemporary therapy while enrolled on prospective multi-institution therapeutic trials that molecular biomarkers identify populations of patients with high-risk disease. Ewing sarcoma is a disease defined by the presence of FET-ETS family fusions and we were successfully able to detect such fusions in 80% of cases. A growing body of literature has suggested that inactivation of *STAG2* and *TP53*, as well as multiple recurrent copy number alterations (CNAs), are associated with poor prognosis in EWS more generally.^26–30^ In the context of localized EWS, we confirm that the presence of recurrent CNAs and pathogenic *TP53* and *STAG2* mutations are individually associated with adverse outcomes, and position *STAG2* mutation as a molecular biomarker for incorporation into future therapeutic trials.

A primary objective of the current study was to identify canonical fusions from newly diagnosed patients treated on prospective multi-institutional trials. Using small amounts of DNA and RNA from archival tissue, we were able to identify canonical Ewing sarcoma FET-ETS family fusions in 80% of cases. While there has been mixed literature on the prognostic significance of various FET-ETS fusions within EWS^31–34^, we found no significant difference in cumulative incidence of relapse for patients with *EWSR1-FLI1* vs. *EWSR1-ERG* fusions, nor did we observe a difference in outcome for patients with Type I vs. other *EWSR1-FLI1* fusions. We identified non-FET-ETS family fusions in a small subset of cases, which included patients with *BCOR-CCNB3*, *EWSR1-ATF1*, and *CIC-NUTM1*, all of which have differential outcomes.^35–37^ Among patients with no fusion identified, we hypothesize that the majority harbor FET-ETS fusions but had inadequate DNA or RNA for sequencing. This result underscores the importance of using a molecular approach to define Ewing sarcoma prospectively from high quality tissue samples for prognostic biomarker research, and by extension, future clinical trials as well.

In 2014, three landscape analyses of Ewing sarcoma genomics identified correlations between alterations in *STAG2* and poor outcomes.^10–12^ Following this work, preclinical and clinical analyses have demonstrated the underlying biologic and clinical impact of *STAG2* deleterious alterations in EWS, further supporting the notion that *STAG2* loss may contribute to the metastatic potential of EWS.^28,38,39^ In our current study of a large patient population enrolled on interventional trials we found that *STAG2* mutation identified the highest risk patients with a cumulative incidence of relapse of 53% (and an EFS of 47%), while patients without a *STAG2* mutation had a cumulative incidence of relapse of 21% (EFS of 76%). Importantly, *STAG2* mutation remained prognostic in a multivariable analysis, supporting the value of this molecular biomarker in identifying localized EWS patients with a higher cumulative incidence of relapse.

Multiple prior studies have suggested a prognostic impact of specific copy number gains as well as overall genomic complexity in EWS more generally.^12,15,16,40^ In our study, we evaluated specific recurrent CNAs in EWS: chromosome 1q gain, 8 gain, 12 gain, or 16q loss. When considered in aggregate, we observed that these recurrent CNAs were strongly associated with a higher cumulative incidence of relapse in localized EWS in univariate analyses. Prior studies have shown a consistent trend towards worse outcomes for patients with *TP53* mutations.^13,14^ In our current study, we observed that patients with *TP53* mutations in their diagnostic tissue have significantly inferior outcomes to those who are *TP53* wild-type in univariate analyses. However, the prognostic value of recurrent CNAs and *TP53* mutations were diminished in multivariable analyses, suggesting that both these molecular features co-occur with other clinical and molecular features of prognostic value in localized EWS.

Prior studies demonstrated that loss of STAG2 protein expression can occur without an identifiable *STAG2* mutation and is associated with poor outcomes in patients with localized EWS.^11,13^ We again identified patients with loss of STAG2 protein expression without an identifiable coding gene mutation in *STAG2*, providing further evidence that patients with EWS lose STAG2 expression through alternative means. We evaluated the prognostic impact of 1) complete STAG2 loss by IHC and 2) the composite biomarker of STAG2 loss by mutation or IHC. Complete STAG2 loss by IHC trended toward an increased risk of relapse and the composite biomarker of STAG2 loss by mutation or IHC was associated with an increased risk of relapse. Our power was limited by the number of cases with available tissue for IHC, and technical challenges related to the use of archival tissue that resulted in staining and slide-related artifacts. Among cases without technical challenges, we observed that STAG2 staining was heterogeneous as has been shown previously, likely due to sub-clonal inactivation. We attempted to take an objective approach to this using H-scores and only calling cases as STAG2 lost if cases had complete loss of staining or a mutation. The true biologic relevance of sub-clonal loss of STAG2 remains unknown. Taken together, these data support STAG2 loss by IHC as an emerging biomarker for identifying patients with localized EWS at high risk of relapse, warranting further technical refinement and future study.

While our study includes a large population of COG patients with localized EWS treated with contemporary therapy and available tissue for molecular sequencing, there were several limitations. The tissue obtained for sequencing included patients treated nearly 20 years prior to initiation of this study, and thus there were cases with limited and old tissue. In our study, we were unable to detect fusions in approximately 20% of patients, even after attempting multiple means of DNA- and RNA-based detection. This was likely due primarily to limitations in tissue quantity and quality but could have also been due to inclusion of fusion-negative small round blue cell tumors. Therefore, cases lacking detectable canonical EWS fusions were excluded from all biomarker evaluations. Tumor size, an important clinical prognostic feature, was missing in a significant proportion of cases in our dataset and was therefore excluded from our multivariable analysis. Despite these limitations, the molecular data for this study were generated from a median number of two unstained slides. These data pave the way for incorporation of molecular biomarkers into prospective clinical trials on which there would be significantly fewer tissue quality and quantity limitations.^41^

In summary, we found that patients with localized EWS and *STAG2* mutations were at the highest risk of relapse. Our findings support the incorporation of *STAG2* mutation and protein expression, *TP53* mutation, and copy number alterations into an integrated molecular biomarker strategy in future clinical trials to inform trial approaches testing treatment intensification and de-escalation. Future work integrating clinical variables into a prognostic analysis may improve the identification of a group of patients at the lowest risk of relapse. Ultimately, integrating clinical and molecular features will enable a further refinement of a risk stratification framework for localized EWS.

## Data Availability

All data produced in the present work are contained in the manuscript.

## PRIOR PRESENTATION

Presented at the Sarcoma Session of the ASCO Annual Meeting, June 2, 2024.

## ACKNOWLEDGEMENTS

This work was supported by COG Biospecimen Bank Grant U24CA196173, the NCI K08 Career Development Award (RG), the DoD PRCRP Fellow Career Development Award (RG), the ASCO Career Development Award (RG), the Rally Foundation Career Development Award (RG), the Boston Children’s Hospital TRP Mentored Career Development Award (RG), the NCI 7U10CA180899-10 Children’s Oncology Group Statistics and Data Center (NJD, MDK, RH), the Pediatric Cancer Research Foundation (KK, JA, BDC), the St. Baldrick’s Foundation, the Children’s Oncology Group Reference Lab Grant, the Pan Mass Challenge Precision for Kids Team (KAJ), and the NCTN Operations Center Grant (U10CA180886).

## AUTHOR CONTRIBUTIONS

**Concept and design**: KAJ, BDC, DSS, RG **Financial support**: KAJ, BDC, DSS, RG **Administrative support**: AW, VB, JS **Collection and assembly of data**: KK, NJD, MDK, RH, AW, VB, JS, RG, DSS, BDC, KAJ, JCS, MT, AJC **Data analysis and interpretation**: NJD, RG, DSS, MDK, RH, KK, AW, VB, JS, CR, BDC, SGD, KAJ, JCS, MT, AJC **Manuscript writing**: RG, DSS, NJD, KK, KAJ, BDC **Final approval of manuscript**: All authors **Accountable for all aspects of the work**: All authors

## DISCLAIMER

The content is solely the responsibility of the authors and does not necessarily represent the official views of the National Institutes of Health.

---

**FIG A1.**
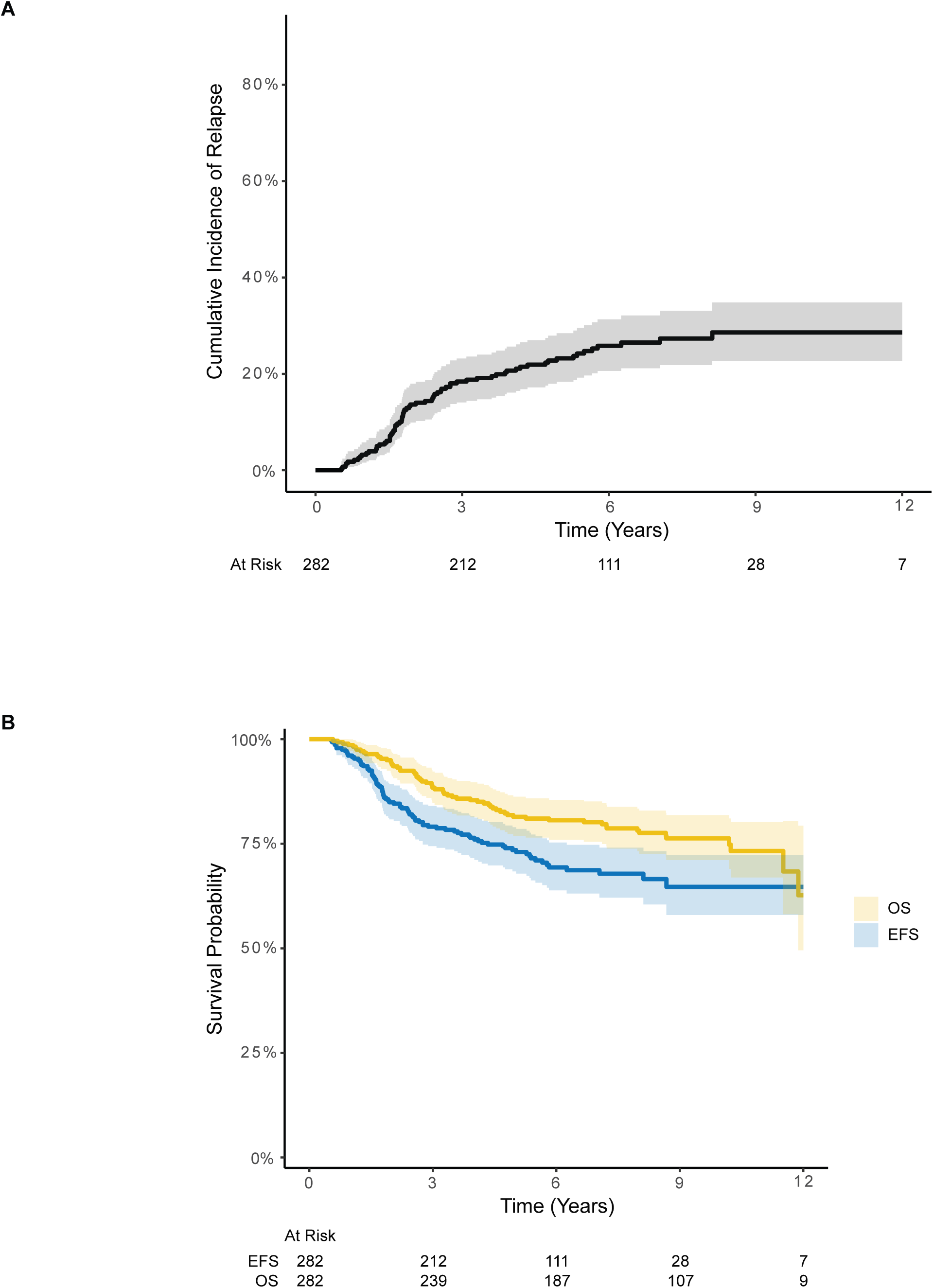
Overall outcomes in analytic cohort of 282 patients with localized EWS. (A) Cumulative incidence of relapse, (B) OS and EFS.

**FIG A2.**
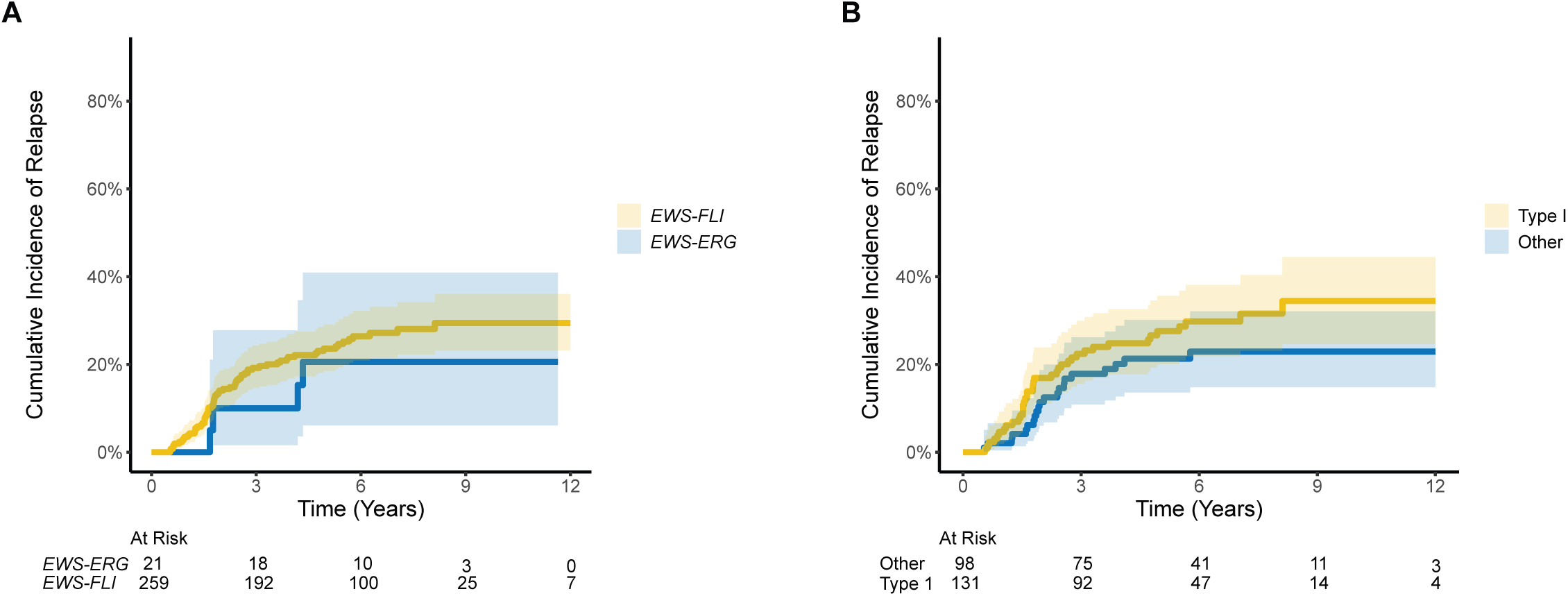
Cumulative incidence of relapse stratified by fusion type. (A) Patients with *EWSR1-FLI1* vs. *EWSR1-ERG* fusions. (B) Patients with *EWSR1-FLI1* Type I vs. all other *EWSR1-FLI1* fusion subtypes.

**FIG A3.**
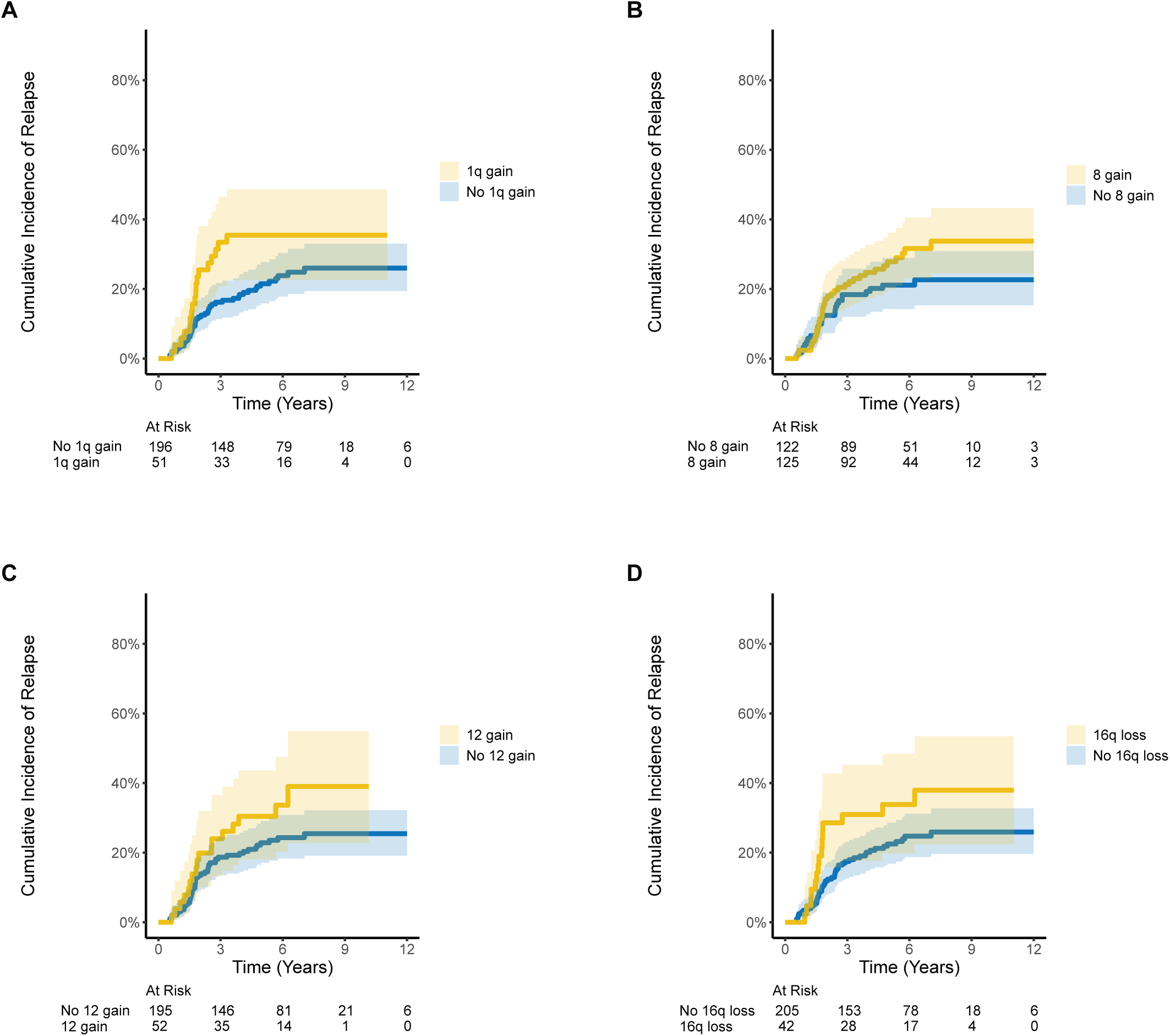
Cumulative incidence of relapse for individual recurrent CNAs. (A) Chromosome 1q gain. (B) Chromosome 8 gain. (C) Chromosome 12 gain. (D) Chromosome 16q loss.

**FIG A4.**
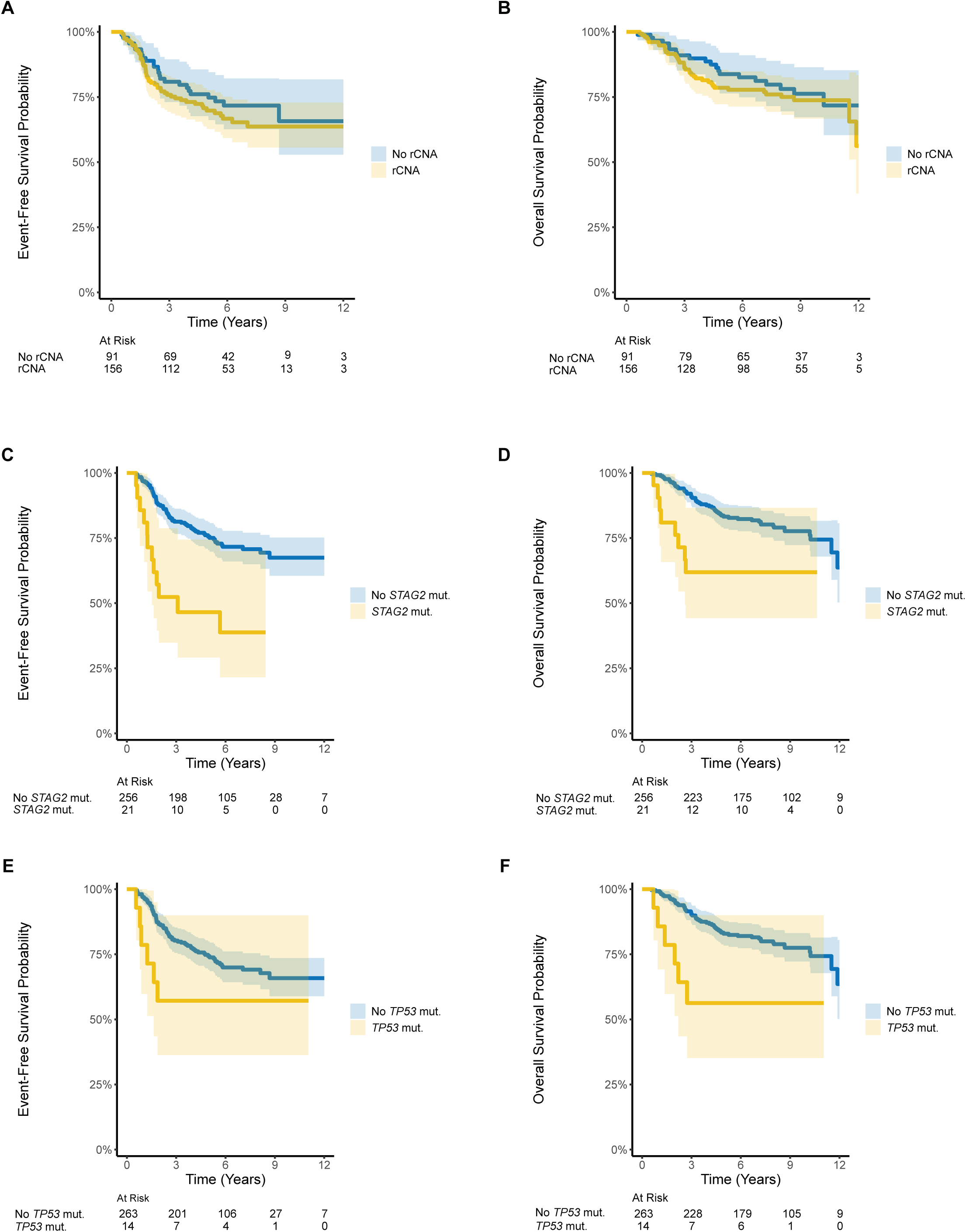
EFS and OS for recurrent CNAs, *STAG2* mutation, and *TP53* mutation. Recurrent CNAs (A) EFS and (B) OS. *STAG2* mutation (C) EFS and (D) OS. *TP53* mutation (E) EFS and (F) OS.

**FIG A5.**
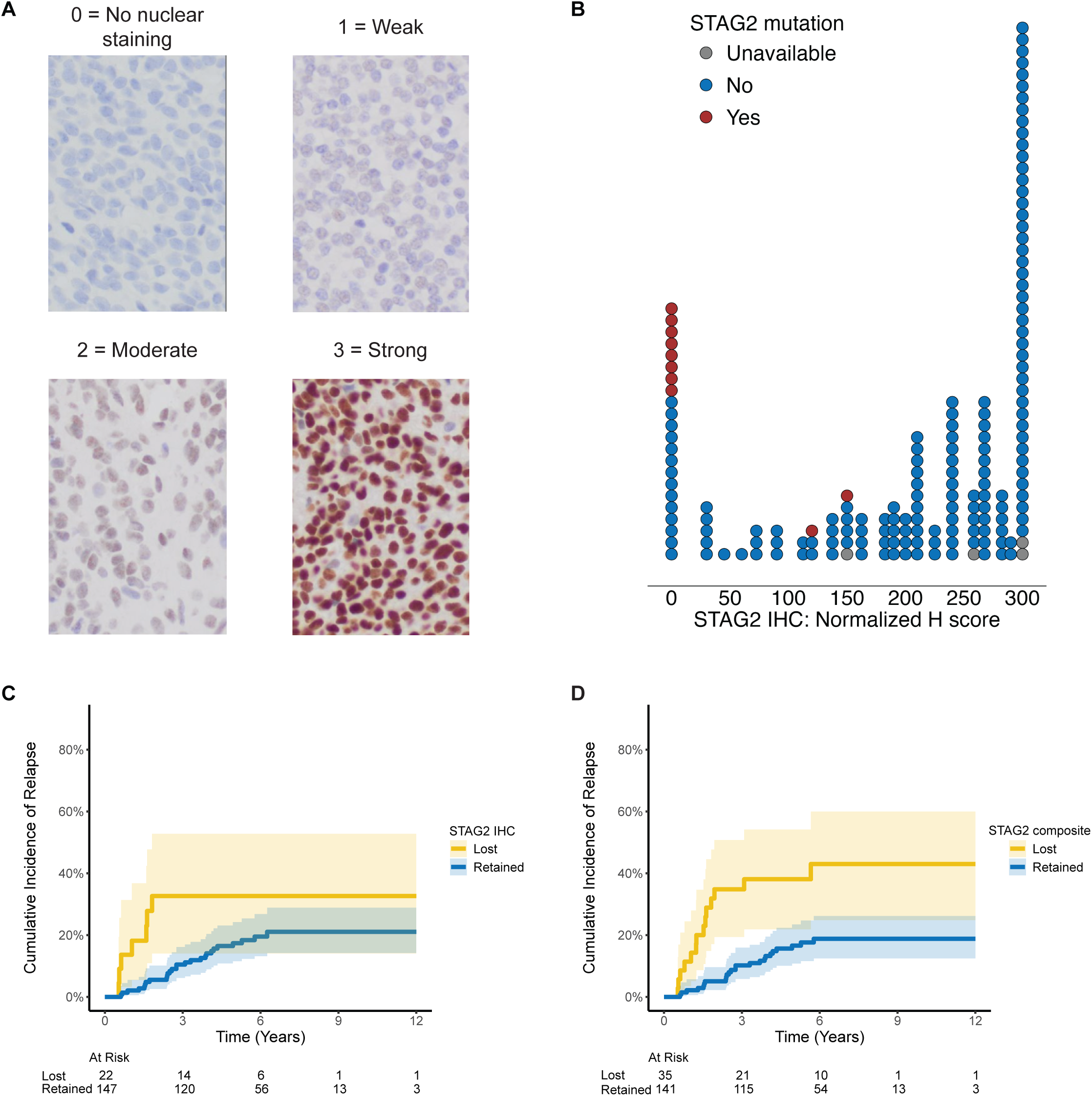
STAG2 immunohistochemistry (IHC). (A) Range of no nuclear staining to strong staining for STAG2 by IHC observed across the analytic cohort. (B) There was high concordance of *STAG2* mutation and complete loss of expression by IHC, and many samples had complete loss in the absence of a detectable *STAG2* mutation. (C) Cumulative incidence of relapse for patients with complete STAG2 loss by IHC vs. those without. (D) Cumulative incidence of relapse for patients with STAG2 loss by mutation or IHC vs. those without.

**TABLE A1.** Genes included on the TranSS-seq assay and LaMPP fusion panel.

| Genes included on custom panels |  |  |  |  |  |
| --- | --- | --- | --- | --- | --- |
| TranSS-seq | LaMPP |  |  |  |  |
| BCOR | ALK | ETV1 | MEAF6 | PPARG | TCF12 |
| CCNB3 | BCOR | ETV4 | MET | PRKACA | TFE3 |
| CIC | BRAF | ETV5 | MKL2 | PRKCA | TFEB |
| DUX4L6 | BRD3 | ETV6 | MYB | PRKCB | TFG |
| DUX4L7 | BRD4 | EWSR1 | MYBL1 | PRKCD | USP6 |
| ERG | C11orf95 | FGFR1 | NCOA1 | QKI | VGLL2 |
| EWSR1 | CAMTA1 | FGFR3 | NCOA2 | RAF1 | YAP1 |
| FLI1 | CCNB3 | FOSB | NTRK1 | RELA | YWHAE |
| FOXO1 | CIC | FOXO1 | NTRK2 | RET |  |
| FUS | CDH11 | FOXO4 | NTRK3 | ROS1 |  |
| PAX3 | DNAJB1 | FUS | NUTM1 | SS18 |  |
| PAX7 | EGFR | GLI1 | PDGFB | SS18L1 |  |
| STAG2 | EPC1 | HMGA2 | PHF1 | STAT6 |  |
| TP53 | ERG | MAML2 | PLAG1 | TAF15 |  |

**TABLE A2.** *A priori* variable selection for multivariable model.

| Variable Considered | Degrees of Freedom Required | Include in Primary MV Model? | Notes |
| --- | --- | --- | --- |
| Age at diagnosis (years) | 1 | Yes | <25% missingness, prior evidence of association with risk of relapse |
| Ethnicity | 1 | No | Not a known prognostic factor associated with risk of relapse |
| Race | 3+ | No | Not a known prognostic factor associated with risk of relapse |
| Sex | 1 | Yes | <25% missingness, prior evidence of association with risk of relapse |
| Primary tumor site | 2 | Yes | <25% missingness, prior evidence of association with risk of relapse |
| Tumor volume (mL) | 1 | No | Prior evidence of association with risk of relapse but >25% missingness and non-uniform measurement on AEWS0031 |
| Interval Compressed Chemotherapy | 1 | Yes | Experimental intervention on AEWS0031, applies to all patients on AEWS1031, proxy for modern therapy regimen |
| Therapy combination | 1 | No | VTC/VDC/IE vs. VDC/IE, showed no significant difference in risk of primary outcome measures per the AEWS1031 primary publication |
| Chemotherapy response (any vs. no viable tumor) | 1 | No | Only collected on the subset of patients who make it to the time point of local control event-free and receive surgery only or surgery followed by radiation therapy, not a known prognostic factor associated with risk of relapse |
| Type of local control | 3 | No | Only collected on the subset of patients who make it to the time point of local control event-free, not a known prognostic factor associated with risk of relapse |
| STAG2 loss by IHC | 1 | No | >25% missingness, investigated in a supplemental analysis |
| EWS Fusion | 3 | No | Not a known prognostic factor associated with risk of relapse, sparse categories outside of EWS-FLI1 |
| TP53 Mutation | 1 | Yes | <25% missingness, primary molecular feature |
| STAG2 Mutation | 1 | Yes | <25% missingness, primary molecular feature |
| 1q gain | 1 | No | Too many terms to support individually with limited dataset, insufficient prior literature demonstrating association with risk of relapse, include as a composite marker indicating whether any copy number change is present |
| 8 gain | 1 | No |  |
| 12 gain | 1 | No |  |
| 16q loss | 1 | No |  |
| rCNA | 1 | Yes | Composite indicator of whether any selected copy number alterations are present |

**TABLE A3.** Multiply imputed multivariable analysis of association of molecular and clinical characteristics with cumulative incidence of relapse.

| Molecular/ clinical characteristic | HR | 95% CI | P value |
| --- | --- | --- | --- |
| rCNA | 1.82 | 0.97, 3.41 | 0.06 |
| TP53 mutation | 1.45 | 0.45, 4.71 | 0.534 |
| STAG2 mutation | 3.42 | 1.69, 6.92 | <0.001 |
| Interval compressed chemotherapy | 0.4 | 0.24, 0.66 | <0.001 |
| Age | 1.12 | 1.07, 1.17 | <0.001 |
| Male sex (relative to female sex) | 0.61 | 0.37, 1.00 | 0.05 |
| Site (relative to non-pelvic) |  |  |  |
| Extraosseous | 0.48 | 0.23, 1.01 | 0.052 |
| Pelvic | 1.01 | 0.57, 1.78 | 0.981 |

## REFERENCES

1 Riggi N, Suvà ML, Stamenkovic I. Ewing’s Sarcoma. N Engl J Med 2021;384:154–64. 10.1056/nejmra2028910.

2 Leavey PJ, Laack NN, Krailo MD, Buxton A, Randall RL, DuBois SG, et al. Phase III Trial Adding Vincristine-Topotecan-Cyclophosphamide to the Initial Treatment of Patients With Nonmetastatic Ewing Sarcoma: A Children’s Oncology Group Report. J Clin Oncol 2021;39:4029–38. 10.1200/jco.21.00358.

3 Caruso J, Shulman DS, DuBois SG. Second malignancies in patients treated for Ewing sarcoma: A systematic review. Pediatr Blood Cancer 2019;66:e27938. 10.1002/pbc.27938.

4 Marina NM, Liu Q, Donaldson SS, Sklar CA, Armstrong GT, Oeffinger KC, et al. Longitudinal follow-up of adult survivors of Ewing sarcoma: A report from the Childhood Cancer Survivor Study. Cancer 2017;123:2551–60. 10.1002/cncr.30627.

5 Dieffenbach BV, Murphy AJ, Liu Q, Ramsey DC, Geiger EJ, Diller LR, et al. Cumulative burden of late, major surgical intervention in survivors of childhood cancer: a report from the Childhood Cancer Survivor Study (CCSS) cohort. Lancet Oncol 2023;24:691–700. 10.1016/s1470-2045(23)00154-7.

6 Chang T-C, Chen W, Qu C, Cheng Z, Hedges D, Elsayed A, et al. Genomic Determinants of Outcome in Acute Lymphoblastic Leukemia. J Clin Oncol 2024;42:3491–503. 10.1200/jco.23.02238.

7 Irwin MS, Naranjo A, Zhang FF, Cohn SL, London WB, Gastier-Foster JM, et al. Revised Neuroblastoma Risk Classification System: A Report From the Children’s Oncology Group. J Clin Oncol 2021;39:3229–41. 10.1200/jco.21.00278.

8 Karski EE, McIlvaine E, Segal MR, Krailo M, Grier HE, Granowetter L, et al. Identification of Discrete Prognostic Groups in Ewing Sarcoma. Pediatr Blood Cancer 2016;63:47–53. 10.1002/pbc.25709.

9 Rodríguez-Galindo C, Liu T, Krasin MJ, Wu J, Billups CA, Daw NC, et al. Analysis of prognostic factors in ewing sarcoma family of tumors. Cancer 2007;110:375–84. 10.1002/cncr.22821.

10 Brohl AS, Solomon DA, Chang W, Wang J, Song Y, Sindiri S, et al. The genomic landscape of the Ewing Sarcoma family of tumors reveals recurrent STAG2 mutation. PLoS Genet 2014;10:e1004475. 10.1371/journal.pgen.1004475.

11 Crompton BD, Stewart C, Taylor-Weiner A, Alexe G, Kurek KC, Calicchio ML, et al. The genomic landscape of pediatric Ewing sarcoma. Cancer Discov 2014;4:1326–41. 10.1158/2159-8290.cd-13-1037.

12 Tirode F, Surdez D, Ma X, Parker M, Deley MCL, Bahrami A, et al. Genomic landscape of Ewing sarcoma defines an aggressive subtype with co-association of STAG2 and TP53 mutations. Cancer Discov 2014;4:1342–53. 10.1158/2159-8290.cd-14-0622.

13 Shulman DS, Chen S, Hall D, Nag A, Thorner AR, Lessnick SL, et al. Adverse prognostic impact of the loss of STAG2 protein expression in patients with newly diagnosed localised Ewing sarcoma: A report from the Children’s Oncology Group. Br J Cancer 2022;127:2220–6. 10.1038/s41416-022-01977-2.

14 Lerman DM, Monument MJ, McIlvaine E, Liu X, Huang D, Monovich L, et al. Tumoral TP53 and/or CDKN2A alterations are not reliable prognostic biomarkers in patients with localized Ewing sarcoma: A report from the Children’s Oncology Group. Pediatr Blood Cancer 2015;62:759–65. 10.1002/pbc.25340.

15 Hattinger CM, Pötschger U, Tarkkanen M, Squire J, Zielenska M, Kiuru-Kuhlefelt S, et al. Prognostic impact of chromosomal aberrations in Ewing tumours. Br J Cancer 2002;86:1763–9. 10.1038/sj.bjc.6600332.

16 Mackintosh C, Ordóñez JL, García-Domínguez DJ, Sevillano V, Llombart-Bosch A, Szuhai K, et al. 1q gain and CDT2 overexpression underlie an aggressive and highly proliferative form of Ewing sarcoma. Oncogene 2012;31:1287–98. 10.1038/onc.2011.317.

17 Shulman DS, Whittle SB, Surdez D, Bailey KM, Alava E de, Yustein JT, et al. An international working group consensus report for the prioritization of molecular biomarkers for Ewing sarcoma. Npj Precis Oncol 2022;6:65. 10.1038/s41698-022-00307-2.

18 Adalsteinsson VA, Ha G, Freeman SS, Choudhury AD, Stover DG, Parsons HA, et al. Scalable whole-exome sequencing of cell-free DNA reveals high concordance with metastatic tumors. Nat Commun 2017;8:1324. 10.1038/s41467-017-00965-y.

19 Klega K, Imamovic-Tuco A, Ha G, Clapp AN, Meyer S, Ward A, et al. Detection of Somatic Structural Variants Enables Quantification and Characterization of Circulating Tumor DNA in Children With Solid Tumors. JCO Precis Oncol 2018;2018:1–13. 10.1200/po.17.00285.

20 Gray RJ. A Class of K-Sample Tests for Comparing the Cumulative Incidence of a Competing Risk. Ann Stat 1988;16:. 10.1214/aos/1176350951.

21 Fine JP, Gray RJ. A Proportional Hazards Model for the Subdistribution of a Competing Risk. J Am Stat Assoc 1999;94:496–509. 10.1080/01621459.1999.10474144.

22 Steyerberg EW. Clinical Prediction Models, A Practical Approach to Development, Validation, and Updating. Stat Biol Heal 2019. 10.1007/978-3-030-16399-0.

23 Jr. FEH. Regression Modeling Strategies, With Applications to Linear Models, Logistic and Ordinal Regression, and Survival Analysis. Springer Ser Stat 2015. 10.1007/978-3-319-19425-7.

24 Buuren S van, Groothuis-Oudshoorn K. mice□: Multivariate Imputation by Chained Equations in R. J Stat Softw 2011;45:. 10.18637/jss.v045.i03.

25 Gamberi G, Cocchi S, Benini S, Magagnoli G, Morandi L, Kreshak J, et al. Molecular Diagnosis in Ewing Family Tumors The Rizzoli Experience—222 Consecutive Cases in Four Years. J Mol Diagn 2011;13:313–24. 10.1016/j.jmoldx.2011.01.004.

26 Tirode F, Consortium for the StJCRHUPCGP and the ICG, Surdez D, Ma X, Parker M, Deley MCL, et al. Genomic Landscape of Ewing Sarcoma Defines an Aggressive Subtype with Co-Association of STAG2 and TP53 Mutations. Cancer Discov 2014;4:1342–53. 10.1158/2159-8290.cd-14-0622.

27 Lerman DM, Monument MJ, McIlvaine E, Liu X, Huang D, Monovich L, et al. Tumoral TP53 and/or CDKN2A alterations are not reliable prognostic biomarkers in patients with localized Ewing sarcoma: A report from the Children’s Oncology Group. Pediatr Blood Cancer 2015;62:759–65. 10.1002/pbc.25340.

28 Shulman DS, Chen S, Hall D, Nag A, Thorner AR, Lessnick SL, et al. Adverse prognostic impact of the loss of STAG2 protein expression in patients with newly diagnosed localised Ewing sarcoma: A report from the Children’s Oncology Group. Br J Cancer 2022;127:2220–6. 10.1038/s41416-022-01977-2.

29 Mackintosh C, Ordóñez JL, García-Domínguez DJ, Sevillano V, Llombart-Bosch A, Szuhai K, et al. 1q gain and CDT2 overexpression underlie an aggressive and highly proliferative form of Ewing sarcoma. Oncogene 2012;31:1287–98. 10.1038/onc.2011.317.

30 Hattinger CM, Pötschger U, Tarkkanen M, Squire J, Zielenska M, Kiuru-Kuhlefelt S, et al. Prognostic impact of chromosomal aberrations in Ewing tumours. Br J Cancer 2002;86:1763–9. 10.1038/sj.bjc.6600332.

31 Zoubek A, Dockhorn-Dworniczak B, Delattre O, Christiansen H, Niggli F, Gatterer-Menz I, et al. Does expression of different EWS chimeric transcripts define clinically distinct risk groups of Ewing tumor patients? J Clin Oncol 1996;14:1245–51. 10.1200/jco.1996.14.4.1245.

32 Alava E de, Kawai A, Healey JH, Fligman I, Meyers PA, Huvos AG, et al. EWS-FLI1 fusion transcript structure is an independent determinant of prognosis in Ewing’s sarcoma. J Clin Oncol 1998;16:1248–55. 10.1200/jco.1998.16.4.1248.

33 Doorninck JA van, Ji L, Schaub B, Shimada H, Wing MR, Krailo MD, et al. Current Treatment Protocols Have Eliminated the Prognostic Advantage of Type 1 Fusions in Ewing Sarcoma: A Report From the Children’s Oncology Group. J Clin Oncol 2010;28:1989–94. 10.1200/jco.2009.24.5845.

34 Deley M-CL, Delattre O, Schaefer K-L, Burchill SA, Koehler G, Hogendoorn PCW, et al. Impact of EWS-ETS Fusion Type on Disease Progression in Ewing’s Sarcoma/Peripheral Primitive Neuroectodermal Tumor: Prospective Results From the Cooperative Euro-E.W.I.N.G. 99 Trial. J Clin Oncol 2010;28:1982–8. 10.1200/jco.2009.23.3585.

35 Palmerini E, Gambarotti M, Italiano A, Nathenson MJ, Ratan R, Dileo P, et al. A global collaboRAtive study of CIC-rearranged, BCOR::CCNB3-rearranged and other ultra-rare unclassified undifferentiated small round cell sarcomas (GRACefUl). Eur J Cancer 2023;183:11–23. 10.1016/j.ejca.2023.01.003.

36 Li A-B, Jiang B-J, Wang H-H, Yang Y-S, Zhang X-B, Lan G-H, et al. Prognostic Factors for Survival in Patients with Clear Cell Sarcoma: An Analysis of the Surveillance, Epidemiology, and End Results (SEER) Database. Méd Sci MonitL: Int Méd J Exp Clin Res 2019;25:6939–45. 10.12659/msm.916705.

37 Gonzaga MI, Grant L, Curtin C, Gootee J, Silberstein P, Voth E. The epidemiology and survivorship of clear cell sarcoma: a National Cancer Database (NCDB) review. J Cancer Res Clin Oncol 2018;144:1711–6. 10.1007/s00432-018-2693-6.

38 Adane B, Alexe G, Seong BKA, Lu D, Hwang EE, Hnisz D, et al. STAG2 loss rewires oncogenic and developmental programs to promote metastasis in Ewing sarcoma. Cancer Cell 2021;39:827–844.e10. 10.1016/j.ccell.2021.05.007.

39 Surdez D, Zaidi S, Grossetête S, Laud-Duval K, Ferre AS, Mous L, et al. STAG2 mutations alter CTCF-anchored loop extrusion, reduce cis-regulatory interactions and EWSR1-FLI1 activity in Ewing sarcoma. Cancer Cell 2021;39:810–826.e9. 10.1016/j.ccell.2021.04.001.

40 Anderson ND, Borja R de, Young MD, Fuligni F, Rosic A, Roberts ND, et al. Rearrangement bursts generate canonical gene fusions in bone and soft tissue tumors. Science 2018;361:. 10.1126/science.aam8419.

41 Reed DR, Grohar P, Rubin E, Binitie O, Krailo M, Davis J, et al. Children’s Oncology Group’s 2023 blueprint for research: Bone tumors. Pediatr Blood Cancer 2023;70:e30583. 10.1002/pbc.30583.

