## Appendix Methods for "Molecular characterization informs prognosis in patients with localized Ewing sarcoma: A report from the Children’s Oncology Group"

**Appendix A**

*Sample Preparation, DNA Extraction, and Sequencing*

Tumor tissue was collected and processed as fresh frozen or FFPE according to institutional protocols and submitted to the Biopathology Center (BPC) at the Children’s Oncology Group (COG). DNA was extracted at the BPC from frozen tumor tissue with the Qiagen Gentra Puregene Tissue kit or from FFPE tumor tissue (two unstained slides) using the Qiagen Allprep FFPE kit. DNA was quantified using PicoGreen, and libraries were prepared using KAPA HyperPrep Kit with Library Amplification (KAPA Biosystems, KK8504) and duplex UMI adapters (IDT).

RNA was extracted at the BPC for a subset of cases using the Roche Highpure hybrid kit or by the Boston Children’s Hospital Laboratory for Molecular Pediatric Pathology (LaMPP) using the Promega Maxwell RSC instrument and reagents or Covaris truXTRAC FFPE total NA Ultra Kit. Nucleic acid quantitation was performed with the Promega Quantus fluorometer using the RNA QuantiFluor kit.

Ultra-low passage whole genome sequencing (ULP-WGS) was performed on sequencing libraries to a target coverage of 0.1x (range <0.01x-0.74x, mean 0.18x). The ichorCNA algorithm was applied on the ULP-WGS BAM files and results were manually curated to determine copy number alterations.^1^

A custom hybrid-capture assay, TranSS-seq,^2^  was run for all samples as previously described, with an anticipated mean target coverage of >150x (range <1x-3896x). The TranSS-seq assay has a validated bait set containing intronic regions of genes commonly involved in sarcoma translocations, including *EWSR1* and *FUS*, as well as exonic regions of the genes *TP53* and *STAG2* (**Appendix Table A1**).

*Identification of Gene Fusions*

Gene fusions were identified using the published algorithms SvABA and BreaKmer.^3,4^ Breakpoint coordinates were determined using UCSC Human BLAT Search with the hg19 assembly. Tumor content was quantified by comparing the translocation reads to wild-type reads with the following formula: % tumor DNA = T/[([W − T]/2) + T], where T is the number of translocation reads and W is the number of wild-type reads.

In the subset of cases with extracted RNA, fusion detection was performed using a custom fusion panel developed by LaMPP (**Appendix Table A1**).  Total nucleic acid or RNA alone was isolated, RNA was converted to cDNA by reverse transcriptase and library preparation was performed using a custom Archer FusionPlex kit (ArcherDX) using anchored multiplex PCR on an Illumina MiSeq sequencer. Sequencing reads were aligned, annotated and analyzed using the Archer Analysis bioinformatics software v6.2.7 system, as previously described.^5^

*EWSR1-FLI1* fusion subtypes were categorized as Type I (defined as including the *EWSR1* transcript to exon 7 or 8 using the canonical transcript NM_005243/ ENST00000397938, and the *FLI1* transcript starting at exon 6 using the canonical transcript NM_002017/ ENST00000527786) or other to enable analysis of outcomes related to fusion subtype.^6^  *EWSR1-FLI1* fusion subtype analysis was restricted to cases with DNA-based translocation calls, unambiguous intronic breakpoints, and only a single fusion subtype detected. Examples of DNA breakpoint data for *EWSR1-FLI1* and *EWSR1-ERG* were visualized using the St. Jude ProteinPaint web application (<https://proteinpaint.stjude.org/>).^7^

*Mutation Identification and Annotation*

*TP53* and *STAG2* mutations were identified using the Mutect2 algorithm in “tumor-only” mode.^8^  Mutations overlapping exons with allele fraction (AF) >= 0.05 and seen in at least three reads were retained as coding variants of possible significance. These candidate mutations were visualized using the Integrative Genomics Viewer (IGV version 2.3.81) and categorized as true positive or false positive calls depending on the depth of sequencing, the number of visualized alternative allele reads, the visualized AF, and the presence of artifacts at or around the examined mutation site. True positive mutations were subsequently curated for evidence of pathogenicity. *TP53* mutations with “Pathogenic” or “Likely Pathogenic” annotations in ClinVar were considered to be pathogenic in the somatic context and included in downstream analyses. *STAG2* mutations with nonsense, frameshift, or high impact splice site variants were considered to be pathogenic in the somatic context and included in downstream analyses. Mutation data for *TP53* and *STAG2* was visualized using the St. Jude ProteinPaint web application (<https://proteinpaint.stjude.org/>).

*Identification of Copy Number Alterations*

Based on prior evidence supporting the relevance of specific copy number alterations (CNAs) in Ewing sarcoma, chromosome 1q gain, chromosome 8 gain, chromosome 12 gain, and chromosome 16q loss were evaluated in this study. CNAs were identified using “.seg” files and manual review of tracings produced during the ichorCNA analysis. Arm level gains and losses were called if greater than or equal to 50% of a chromosomal arm was altered. We assessed the relationship between each CNA and outcome separately and as a composite molecular feature of any of the four evaluated CNAs (collectively referred to as recurrent CNAs).

*STAG2 Immunohistochemistry*

Immunohistochemical staining was performed for STAG2 using the mouse anti-human monoclonal antibody (Santa Cruz SA-2 (J-12): sc-81852).^9^

Two pediatric pathologists evaluated intensity of nuclear staining across the total proportion of tumor cells visualized using the H-score, calculated as follows: (1 × percentage of weak staining) + (2 × percentage of moderate staining) + (3 × percentage of strong staining) within the tumor cells, ranging from 0 to 300. Due to variability in the staining of controls (endothelial cells), the H-score was subsequently normalized by comparing to maximum staining from the internal positive control, and capped at 300, as follows: Normalized H score = min{300, (H score)*3/(Highest staining from the internal positive control)}. Cases with inadequate viable tumor cells, extensive necrosis, features of harsh acid decalcification, or with no internal controls with observable STAG2 expression were considered uninterpretable.

Samples with normalized H scores of 0 were determined to have “complete loss” of STAG2 expression, whereas samples with scores > 0 were considered to have at least partially retained expression. Samples determined to have complete loss of STAG2 expression were combined with those identified as carrying loss-of-function *STAG2* mutations for the composite biomarker “STAG2 loss by mutation or IHC.”

**References**

1. Adalsteinsson VA, Ha G, Freeman SS, et al. Scalable whole-exome sequencing of cell-free DNA reveals high concordance with metastatic tumors. *Nat Commun*. 2017;8(1):1324.

2. Klega K, Imamovic-Tuco A, Ha G, et al. Detection of Somatic Structural Variants Enables Quantification and Characterization of Circulating Tumor DNA in Children With Solid Tumors. *JCO Precis Oncol*. 2018;2018(2):1-13.

3. Abo RP, Ducar M, Garcia EP, et al. BreaKmer: detection of structural variation in targeted massively parallel sequencing data using kmers. *Nucleic Acids Res*. 2015;43(3):e19-e19.

4. Wala JA, Bandopadhayay P, Greenwald NF, et al. SvABA: genome-wide detection of structural variants and indels by local assembly. *Genome Res*. 2018;28(4):581-591.

5. Fisch AS, Church AJ. Special Considerations in the Molecular Diagnostics of Pediatric Neoplasms. *Clin Lab Med*. 2022;42(Nature Medicine 2022):349-365.

6. Gamberi G, Cocchi S, Benini S, et al. Molecular Diagnosis in Ewing Family Tumors The Rizzoli Experience—222 Consecutive Cases in Four Years. *J Mol Diagn*. 2011;13(3):313-324.

7. Zhou X, Edmonson MN, Wilkinson MR, et al. Exploring genomic alteration in pediatric cancer using ProteinPaint. *Nat Genet*. 2016;48(1):4-6.

8. Cibulskis K, Lawrence MS, Carter SL, et al. Sensitive detection of somatic point mutations in impure and heterogeneous cancer samples. *Nat Biotechnol*. 2013;31(3):213-219.

9. Shulman DS, Chen S, Hall D, et al. Adverse prognostic impact of the loss of STAG2 protein expression in patients with newly diagnosed localised Ewing sarcoma: A report from the Children’s Oncology Group. *Br J Cancer*. 2022;127(12):2220-2226.
